# CLEC10A marks an MHC class II-high tumour myeloid state that lies in T-cell-rich neighbourhoods, and its survival association is seen in MYCN non-amplified neuroblastoma

**DOI:** 10.64898/2026.10.30.26366893

**Authors:** Shoryu Takayama, Keisuke Yamazaki, Tomonori Tsuchiya, Hiroki Kanamori, Atsuki Naoe, Yasuhiro Kondo, Shunsuke Watanabe, Toshihiro Yasui, Eri Ogawa, Mikihiro Inoue

## Abstract

**Background:** CLEC10A (CD301) is widely used as a marker of CD1c-positive conventional type 2 dendritic cells (cDC2). Whether it carries prognostic information in neuroblastoma has not been examined, and what it marks inside tumour myeloid cells has not been separated from the artefacts that dominate marker-positive versus marker-negative comparisons.

**Methods:** CLEC10A and overall survival were analysed in three neuroblastoma cohorts (920 tumours) using MYCN-stratified Cox models and random-effects meta-analysis, with a Knapp–Hartung sensitivity analysis. In the GSE147766 immune-cell atlas, CLEC10A-positive cells were matched to negative cells on sequencing depth and myeloid subtype within patient before patient-paired pseudobulk differential expression. In six Visium HD sections (8 µm bins) from five patients, T-cell transcript density within 48 µm of matched bins was compared with patient-level inference and a spatial block bootstrap.

**Results:** Detection of CLEC10A rose 2.48-fold per standard deviation of log sequencing depth, and only 18% of CLEC10A-positive cells were cDC2-like. After matching, 107 genes remained differentially expressed; MHC class II dominated, reproducing within monocytes and macrophages alone (7 of 7 patients), while the S100 alarmin programme was lower. Decomposing the design showed the apparent MYC-target enrichment to be attributable to depth, and the MHC class II signal invariant to every correction. CLEC10A-positive myeloid bins occupied T-cell-richer neighbourhoods in all five patients (pooled paired difference 0.051, 95% bootstrap interval 0.003 to 0.134). Survival association was present in MYCN non-amplified disease (pooled hazard ratio 0.77) and absent in amplified disease, a difference that three cohorts cannot establish.

**Conclusions:** CLEC10A marks an MHC class II-high, alarmin-low myeloid state rather than cDC2 abundance, and that state lies near T cells. Whether these cells prime T cells is untested.

## Introduction

Neuroblastoma is the commonest extracranial solid tumour of childhood and the most clinically heterogeneous. Tumours in infants regress spontaneously or mature without cytotoxic treatment, while metastatic disease in older children survives intensive multimodal therapy in only about half of cases. Age, stage and tumour biology — above all amplification of *MYCN* — dominate outcome and are formalised in the risk classifications that now direct treatment [1–3].

That heterogeneity extends to the immune compartment, and the therapeutic stakes are not hypothetical. Antibody against the disialoganglioside GD2, combined with cytokines and isotretinoin, improved event-free survival in high-risk disease and is standard care [4]; GD2-directed chimeric antigen receptor T cells have since produced durable responses in relapsed and refractory disease [5]. Both act on the tumour cell surface. Approaches that depend on an endogenous T-cell response have fared less well: in the paediatric KEYNOTE-051 trial, objective responses to pembrolizumab among neuroblastoma patients were rare [6]. Two features of the disease help explain that. Neuroblastoma carries one of the lowest somatic mutation burdens of any human cancer, so the supply of neoantigens is small [7]; and MHC class I expression on neuroblastoma cells is low, a defect linked to *MYCN* amplification in work that long predates the modern immunotherapy era [8]. A tumour that presents little through class I is a difficult target for CD8 T cells, which places more weight on what the antigen-presenting compartment of the microenvironment is doing.

What is known of that microenvironment is consistent and, in places, specific. Lymphocytes outnumber myeloid cells in the immune compartment, yet it is the myeloid side that has carried the clearest prognostic signal: the strength of the inflammatory and macrophage-associated gene programme in metastatic tumours predicts outcome beyond established markers, most clearly in *MYCN* non-amplified disease [9]. *MYCN*-amplified tumours are repeatedly described as immunologically cold, with lower expression of antigen-presentation and interferon-associated programmes [10,11]. Where cytotoxic lymphocytes are present they matter: T-cell infiltration and clonal T-cell receptor expansion track favourable outcome in *MYCN* non-amplified and therapy-resistant disease [12,13]. Single-cell atlases of the neuroblastoma immune compartment have since made this description cellular rather than bulk [14,15], and they confirm that mononuclear phagocytes, not dendritic cells, make up most of it.

The dendritic-cell fraction is nevertheless the part that determines whether a T-cell response can be initiated or sustained, since the density and functional state of intratumoural dendritic cells constrain how effectively any T-cell-directed therapy can work [16]. Attention has centred on type-1 conventional dendritic cells, which cross-present to CD8 T cells; but type-2 conventional dendritic cells (cDC2) and the MHC class II-restricted CD4 help they provide can by themselves drive protective antitumour immunity [17]. In a tumour whose class I presentation is impaired, that route is of more than academic interest. It is also the route about which least is known in neuroblastoma, because the antigen-presenting fraction is small and because most analyses have measured how many such cells are present rather than what state they are in.

CLEC10A (CD301, MGL) sits at that junction, and is where this study begins. It encodes a C-type lectin whose ligand is terminal N-acetylgalactosamine, including the tumour-associated Tn antigen [18]; it is enriched in human CD1c-positive cDC2 and is routinely treated as a cDC2 identity marker [19,20], including in the harmonised neuroblastoma reference atlas, which calls cDC2 by CD1C, CD1E and CLEC10A [14]. Higher CLEC10A in a bulk transcriptome has accordingly been read as a larger cDC2 compartment. In other tumour types that reading has been taken a step further: in lung adenocarcinoma, higher CLEC10A has been reported to accompany greater immune infiltration and better outcome [21]. In neuroblastoma it has not been tested, and what such an association would mean has not been asked. The cDC2-abundance reading also sits awkwardly with what is now known of the mononuclear phagocyte system, in which transcriptional programmes are shared across monocytes, macrophages and dendritic cells rather than partitioned cleanly between them [22].

We had three aims. The first was to ask whether CLEC10A expression carries prognostic information in neuroblastoma at all and, where cohorts disagree, whether that disagreement reflects genuine non-replication or differences in cohort composition. The second was to identify what transcriptional state, if any, CLEC10A marks inside tumour myeloid cells once depth and composition are controlled. The third was to ask where in the tissue that state sits relative to T cells. We addressed the first with prespecified *MYCN*-stratified analysis and cross-cohort meta-analysis in three bulk cohorts [23–25], the second by re-analysing the GSE147766 neuroblastoma immune-cell atlas [15] with an explicit matched design, and the third in Visium HD sections at 8 µm resolution [26].

## Methods

### Study design and data sources

Three questions were fixed before analysis: whether CLEC10A expression was associated with overall survival; whether that association differed by MYCN status; and which myeloid population and transcriptional state carried CLEC10A. A fourth question — whether CLEC10A-positive myeloid cells occupy T-cell-rich neighbourhoods — was specified after the single-cell result and is reported as such. Bulk primary-tumour cohorts were GSE85047 [23], GSE62564 [24] and TARGET-NBL [25]. The single-cell dataset was GSE147766 [15].

The spatial dataset was GSE332618 [26]. Analyses were restricted to samples with the required expression, clinical and outcome fields. This study used only publicly available, de-identified data and did not require institutional review board approval.

### Bulk expression and survival analysis

Gene identifiers and clinical records were harmonised within each cohort, retaining one primary tumour sample per patient in TARGET. CLEC10A was standardised within cohort. Kaplan–Meier curves used a within-cohort median split for display only and are shown per cohort in Supplementary Fig. S1A–C; the hazard ratio across dichotomisation thresholds from the 25th to the 75th percentile is given in Supplementary Fig. S1D. Cox models treated CLEC10A as a continuous variable per 1-SD increase. Adjusted models included the available covariates age at diagnosis ≥18 months, stage 4 and MYCN amplification; high-risk status was structurally collinear and produced an unstable fit in GSE62564, so the prespecified primary adjusted model omitted it and the full model was retained as a sensitivity analysis (Supplementary Fig. S1). The same covariate set was used in all three cohorts; the estimates entering Fig. 1A and the pooled adjusted estimate come from that model in every cohort. Cut-off sensitivity, the full covariate-adjustment ladder, scaled Schoenfeld residual tests of the proportional-hazards assumption [27] and follow-up horizon sensitivity are reported in Supplementary Fig. S1. Ridge-penalised versions of the fully adjusted models are provided as source data.

**Figure 1.**
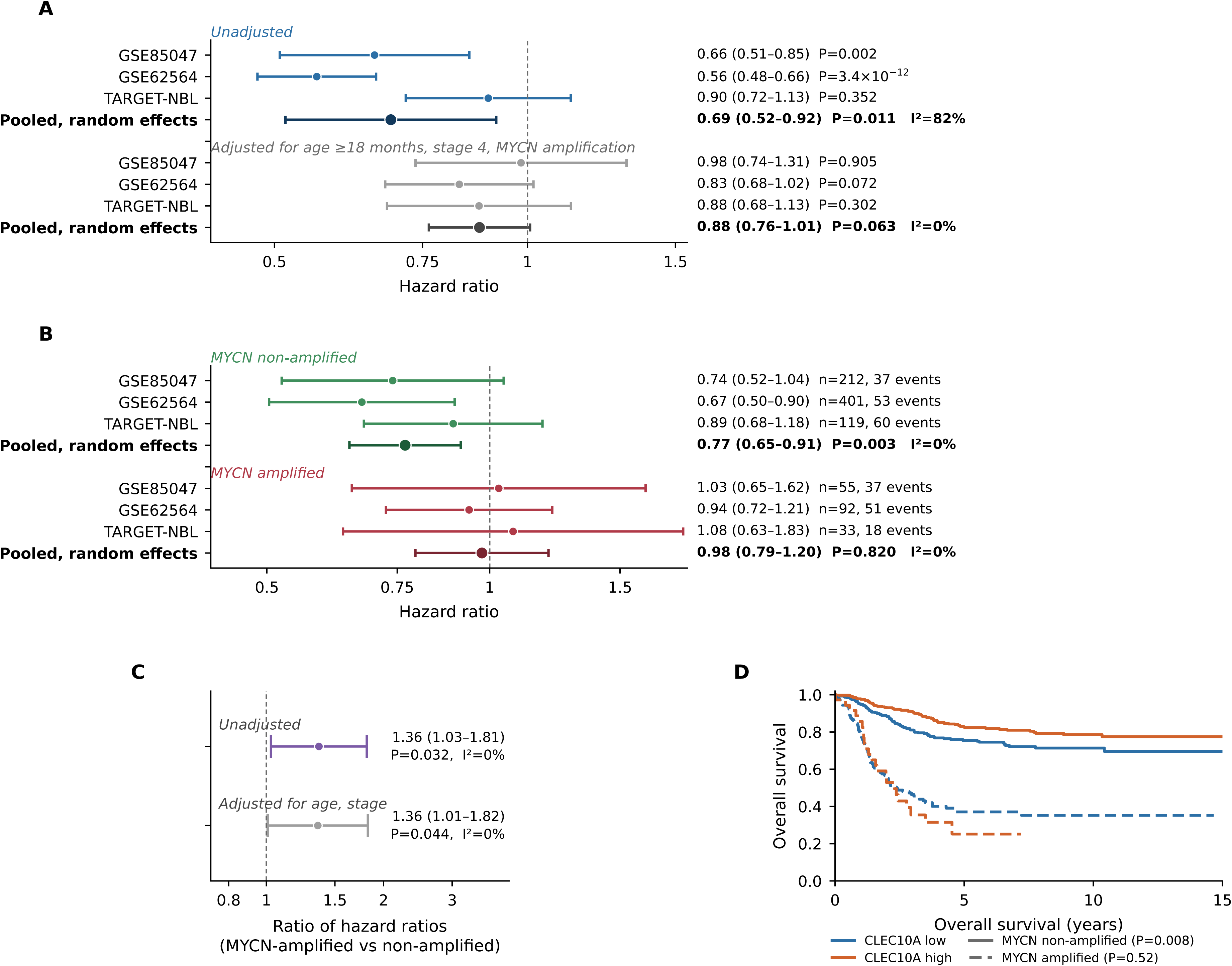
CLEC10A expression and overall survival in three neuroblastoma cohorts, overall and within *MYCN* strata. **A**, Cox hazard ratios for continuous CLEC10A per 1-SD higher within-cohort expression, unadjusted and adjusted for age ≥18 months, stage 4 and *MYCN* amplification, with DerSimonian–Laird random-effects pooled estimates and I². The same covariate set is used in all three cohorts, and the null result in TARGET-NBL is shown without exclusion. **B**, The same estimates within each *MYCN* stratum. The between-cohort heterogeneity seen overall (I²=82%, **A**) is absent within either stratum. **C**, Random-effects meta-analysis of the three within-cohort CLEC10A × *MYCN* interaction terms, expressed as a ratio of hazard ratios, that is, the multiplicative change in the CLEC10A hazard ratio in *MYCN*-amplified relative to non-amplified tumours. **D**, Pooled overall survival by within-cohort median split, with *MYCN* status shown by line style; log-rank P per stratum. Confidence bands are omitted for legibility and the corresponding intervals are given in **B**. Cohort composition is given in Table 1, and the per-cohort survival curves and sensitivity analyses in Supplementary Figure S1. These intervals treat a between-study variance estimated from three studies as known; Supplementary Table S9 gives the Knapp–Hartung and leave-one-cohort-out results. CI, confidence interval.

**Table 1.** Characteristics of the three neuroblastoma cohorts.

| Characteristic | GSE85047 | GSE62564 | TARGET-NBL |
|---|---|---|---|
| Patients, n | 269 | 498 | 153 |
| Deaths, n (%) | 75 (28) | 105 (21) | 79 (52) |
| Median follow-up of survivors, years | 5.3 | 6.5 | 9.1 |
| Maximum follow-up, years | 17.7 | 18.1 | 14.7 |
| Age ≥18 months, n (%) | 128 (48) | 193 (39) | 124 (81) |
| Stage 4, n (%) | 123 (46) | 183 (37) | 126 (82) |
| MYCN amplified, n (%)<sup>a</sup> | 55 (21) | 92 (19) | 33 (22) |
| High risk, n (%)<sup>b</sup> | not recorded | 239 (48) | 127 (83) |
| MYCN non-amplified, n (deaths) | 212 (37) | 401 (53) | 119 (60) |
| MYCN amplified, n (deaths) | 55 (37) | 92 (51) | 33 (18) |

<sup>a</sup>MYCN status was missing for 2 patients in GSE85047, 5 in GSE62564 and 1 in TARGET-NBL; percentages are of patients with a recorded status. <sup>b</sup>Risk group was not available in GSE85047 and was therefore excluded from the primary adjusted model (Methods).

### Cross-cohort meta-analysis and interaction testing

Cohort-specific log hazard ratios and standard errors were combined by DerSimonian– Laird random-effects meta-analysis [28]; heterogeneity is reported as I². The same procedure was applied within each MYCN stratum. Because three studies cannot support a reliable estimate of the between-study variance, every pooled estimate was recomputed with restricted maximum likelihood in place of DerSimonian–Laird, and with the Knapp– Hartung variance estimator, which rescales the variance by the weighted residual mean square and uses a t quantile on k−1 degrees of freedom; the modified form, which does not allow that rescaling to shrink the variance below its fixed-effect value, is reported alongside. At k=3 the t quantile is 4.303, 2.2 times the normal quantile, so the comparison measures directly what the normal approximation assumes. Each pooled estimate was also recomputed omitting one cohort at a time. The full comparison is Supplementary Table S9. The CLEC10A × MYCN interaction was estimated within each cohort from a Cox model containing continuous CLEC10A, MYCN status and their product, with and without age and stage covariates, and the three interaction coefficients were then meta-analysed. The pooled interaction is reported as a ratio of hazard ratios, that is, the multiplicative change in the CLEC10A hazard ratio in MYCN-amplified relative to non-amplified tumours.

### Immune-context contrasts

Gene-level amplified-versus-non-amplified contrasts used within-cohort standardised expression in linear models adjusted for age and stage, followed by robust empirical-Bayes estimation and random-effects meta-analysis with restricted maximum likelihood. Three immune modules were fixed without reference to survival. The APC module comprised CLEC10A, CD1C and FCER1A. The antigen-presentation module comprised CIITA, CD74, HLA-DRA, HLA-DRB1, HLA-DPB1, TAP1, PSMB8, PSMB9 and HLA-DPA1. The T-cell-inflamed module comprised CXCL9, CXCL10, STAT1, ISG15, IFIT1, IFIT2, IFIT3, CD8A, CD8B, TRAC, CCL5 and GZMA. Scores were means of available within-cohort gene z scores and were then standardised within cohort.

### Single-cell processing and scope

Counts for each GSE147766 sample were converted to a common gene space (32,966 genes shared by all samples) and the published myeloid/APC annotation and UMAP embedding were used unchanged [15]; CLEC10A was not a variable feature and was not used to define annotations. NB12 and NB18 were treated as one biological patient according to the source metadata, giving 8,219 cells from 17 biological patients. MYCN status was mapped from the original study’s Table S1. The primary analysis was restricted to MYCN non-amplified tumours (8,092 cells, 14 biological patients), because MYCN-amplified tumours contributed only 127 APCs from 3 patients. The identical pipeline was also run on all patients as a scope-sensitivity analysis (Supplementary Fig. S5); because the amplified tumours add only one further eligible patient, this is a sensitivity check rather than an independent replication. CLEC10A positivity was defined as at least one raw UMI.

### Control of sequencing depth and composition

Two confounders were quantified before any differential expression was run. Per-cell depth was compared between CLEC10A-positive and negative cells within each patient (paired Wilcoxon test on patient medians) and modelled by within-patient logistic regression of CLEC10A positivity on standardised log total UMI. Subtype composition of the CLEC10A-positive and negative pools was tabulated.

Both were then controlled by matching. Within each combination of biological patient and myeloid subtype, each CLEC10A-positive cell was matched without replacement to the nearest unmatched CLEC10A-negative cell on log total UMI, using a calliper of 0.20 SD of log UMI across the analysed cells. Because matching was performed inside patient × subtype strata, the matched positive and negative sets have identical subtype composition by construction. Balance was verified by comparing depth distributions after matching; the assignments are given in Supplementary Table S6.

Matching was used in place of covariate adjustment because the marker defining the compared groups is itself detection-based. Whether a transcript is observed in a droplet library is governed by sampling at the depth achieved [29], so depth here acts on the exposure rather than on the outcome, and adjustment for the cellular detection rate of the kind MAST performs [30] cannot remove confounding from a contrast already formed on a depth-dependent split. Matching equalises the compared sets on depth before the contrast is formed, which is the standard remedy when a measured covariate drives assignment to the compared groups [31,32]; performing it inside patient × subtype strata removes the compositional confounder at the same time, and aggregating to biological patients before testing removes the pseudoreplication that inflates single-cell differential expression when cells rather than donors are treated as independent units [33,34]. Naive and matched contrasts are reported side by side throughout.

Because matching changes depth, composition and the number of cells together, the same patient-paired model was run under five designs on the same cells in order to separate them: unmatched; negatives reduced at random to the number of positives within patient; matched on log total UMI only; matched on myeloid subtype only; and matched on both. Randomised designs were repeated over five seeds. Each run was scored by the number of genes at FDR<0.05 and, for Hallmark MYC targets v1 and the MHC class II module, by a competitive rank test of the member genes’ Wald statistics against all other tested genes, reported as a signed −log₁₀ P so that every design is scored identically. Results are in Supplementary Table S11, with the individual runs in Supplementary Table S11b.

### Differential expression and pathway analysis

Raw counts of matched cells were aggregated by biological patient and CLEC10A status. A patient contributed a pair when at least 10 matched cells were present in each group. Aggregating to biological patients before testing removes cell-level pseudoreplication [33,34]. Differential expression used DESeq2 [35] with the design ∼patient + CLEC10A status, restricted to genes with at least 5 counts in at least half the pseudobulk libraries, with Benjamini–Hochberg control of the false-discovery rate [36]. CLEC10A itself was excluded from all reported results and from the ranked list, because it defines the contrast; it is shown only as a positive control in Supplementary Fig. S3. Stability was assessed by omitting each patient in turn and re-testing the significant gene set.

Genes were ranked by the DESeq2 Wald statistic and tested by pre-ranked gene-set enrichment analysis [37] against the Hallmark [38] and GO Biological Process collections, with gene sets of 15–500 represented genes and 1,000 permutations. Because ribosomal protein transcripts dominate depth-related signal, all analyses were repeated after removing RPL, RPS, RPLP, MRPL, MRPS and mitochondrial genes, and both versions are reported. Two prespecified module scores were also computed per cell as the summed counts of their member genes divided by total counts: an MHC class II module (HLA-DRA, HLA-DRB1, HLA-DPA1, HLA-DPB1, HLA-DQA1, HLA-DQB1, HLA-DMA, HLA-DMB, CD74, CIITA) and an S100/inflammatory monocyte module (S100A8, S100A9, S100A12, FCN1, VCAN, SERPINA1, CD55, FCGR3A, STXBP2, CCL4, IL1B, CXCL8) [39,40]. Patient means were compared by paired Wilcoxon test.

### Spatial analysis

GSE332618 comprises 16 Visium HD sections (Space Ranger 4.0.1, 8 µm bins, 18,132-gene probe panel). The platform reads a probe panel from a continuous 2 µm barcode lattice that is binned for analysis; at 8 µm a bin holds one to a few cells, and immune populations have been resolved at this scale in other tumour types [41]. Six xenograft sections were excluded because they contain no human immune compartment, leaving 10 patient sections. Sections were further required to carry sufficient immune content for a myeloid contrast, defined before any outcome was examined as at least 10% of in-tissue bins positive for a myeloid marker and at least 30 CLEC10A-positive myeloid bins; six sections from five patients met both criteria (one patient contributed two sections). Per-section bin counts and immune content are given in Supplementary Table S7.

Bins outside tissue were removed. A bin was called positive for a programme when the summed raw counts of its member genes were at least one. The myeloid set was LYZ, AIF1, CTSS, TYROBP, FCER1G, C1QA, C1QB, C1QC and CD68; the T-cell set was CD3D, CD3E, CD3G, CD2, TRAC and TRBC2; the tumour set was PHOX2B, TH, DBH, CHGA, CHGB, HAND2, GATA3 and DLK1.

Two properties of these data make a detection-based spatial outcome unsafe, and both were quantified first. The probability that a bin was called T-cell positive rose with bin sequencing depth in five of the six sections, the ratio between the highest and the lowest depth quintile ranging from 0.97 to 10.1 across sections, although the shape of the relationship differed between sections and was not monotone in all of them (Fig. 3B); and bin depth was strongly spatially autocorrelated, the depth of a bin predicting the mean depth of its surrounding ring at Spearman ρ 0.783 to 0.891 for the 48 µm ring over which the outcome is computed, and 0.871 to 0.942 for a 24 µm ring. A deeper bin therefore sits in a deeper neighbourhood, in which T cells are more likely to be *detected* irrespective of whether they are more likely to be *present*. Distance to the nearest T-cell-positive bin is consequently depth-driven, and on an 8 µm grid it is also coarsely discretised. We therefore defined the outcome as the number of T-cell transcripts divided by the total UMI within a 48 µm radius of each bin, excluding the bin itself, expressed per 1,000 UMI. Normalising by local UMI removes the direct effect of library size on the outcome. It does not remove every path: the denominator also depends on local cell density and on the tumour, stromal and immune composition of the neighbourhood, so the ratio can move for reasons other than the number of T cells present.

Each CLEC10A-positive myeloid bin was then matched, without replacement and within section, to a CLEC10A-negative myeloid bin with the same myeloid count (capped at six) and the same log total UMI rounded to one decimal place. Matching was verified to equalise not only the bin’s own depth but the depth of its neighbourhood (ratio of mean neighbourhood UMI, CLEC10A-positive to matched negative, 0.963 to 1.003 across sections). Matched pairs were compared within section as paired differences (assignments in Supplementary Table S8), summarised by the Hodges–Lehmann estimator. Two dependencies rule out treating bins or sections as independent. Neighbourhoods of 48 µm overlap, so between 70% and 100% of analysed bins per section lie within 96 µm of another analysed bin; and two of the six sections come from one patient, leaving five independent patients. Rank tests on so few units are also bounded: with five units the smallest attainable two-sided exact P is 0.0625, so such a test cannot reach conventional significance however large the effect.

Uncertainty was therefore obtained by a hierarchical bootstrap with 4,000 draws. Patients were resampled with replacement; within each sampled patient’s section or sections, contiguous square tiles of bins were resampled with replacement and the paired statistic recomputed, so that local spatial structure is preserved rather than broken by exchanging individual bins. Tiles of 25 bins (200 µm) were used, with 12 and 50 bins as sensitivity analyses. The per-patient effect is the mean over that patient’s sections, and the reported estimate is the mean over patients with a percentile interval. A naive bootstrap over pairs is reported alongside, to show what ignoring the spatial dependence would have claimed. The analysis was repeated omitting each patient in turn (Supplementary Table S10). Unmatched and matched results are reported together, as in the single-cell analysis.

MYCN amplification status is not available in the GSE332618 sample annotation, so the spatial analysis could not be stratified by MYCN and is reported without stratification.

### Statistics

Tests were two-sided. The unmatched contrast is reported alongside the matched contrast throughout so that the effect of each correction is visible. No P value is used in place of an effect estimate, confidence interval or non-estimable result.

## Results

### CLEC10A is associated with survival in two of three cohorts, and the discrepancy is explained by MYCN status

Cohort characteristics are summarised in Table 1. Overall-survival analysis included 269 tumours and 75 deaths in GSE85047, 498 tumours and 105 deaths in GSE62564, and 153 tumours and 79 deaths in TARGET-NBL. Median-split curves separated in GSE85047 (log-rank P=0.013) and GSE62564 (P<0.001) but not TARGET-NBL (P=0.557; Supplementary Fig. S1A–C).

Continuous models agreed. Each 1-SD increase in CLEC10A was associated with lower mortality in GSE85047 (HR 0.66, 95% CI 0.51–0.85, P=0.0016) and GSE62564 (HR 0.56, 0.48–0.66, P=3.4×10⁻¹²) but not in TARGET-NBL (HR 0.90, 0.72–1.13, P=0.35). The pooled random-effects estimate was 0.69 (0.52–0.92, P=0.011), but with substantial heterogeneity (I²=82%; Fig. 1A). After adjustment for age, stage and MYCN status the estimates attenuated to 0.98 (0.74–1.31), 0.83 (0.68–1.02) and 0.88 (0.68–1.13), pooling to 0.88 (0.76–1.01, P=0.063, I²=0%). CLEC10A is therefore not an independent prognostic variable.

The three cohorts differ markedly in composition (Table 1). TARGET-NBL comprises 82% stage 4 tumours, 81% patients aged ≥18 months and 83% high-risk disease, against 37%, 39% and 48% in GSE62564, and 52% of TARGET patients died versus 21% in GSE62564. This raised the possibility that the discrepancy reflects who is in each cohort rather than whether the association is real.

Stratifying by MYCN status resolved the discrepancy (Fig. 1B). In MYCN non-amplified tumours the hazard ratio was 0.74 (0.52–1.04) in GSE85047, 0.67 (0.50–0.90) in GSE62564 and 0.89 (0.68–1.18) in TARGET-NBL, pooling to 0.77 (0.65–0.91, P=0.003) with no detectable heterogeneity (I²=0%). In MYCN-amplified tumours the corresponding estimates were 1.03 (0.65–1.62), 0.94 (0.72–1.21) and 1.08 (0.63–1.83), pooling to 0.98 (0.79–1.20, P=0.82, I²=0%). The heterogeneity present in the overall analysis (I²=82%) was thus no longer detectable after stratification, and TARGET-NBL — the cohort in which the association had appeared to fail — behaved like the others once its MYCN non-amplified patients were considered separately.

These pooled intervals should not be read as firmly as their width suggests. The between-study variance was estimated as zero in every stratum, so the random-effects estimate is numerically the fixed-effect estimate, and the normal-based interval treats a variance estimated from three studies as if it were known. Substituting the modified Knapp–Hartung estimator, which uses a t quantile on two degrees of freedom and does not allow the variance to shrink below its fixed-effect value, leaves the point estimates unchanged and widens the intervals substantially: 0.77 (0.53–1.12, P=0.097) in MYCN non-amplified tumours, 0.98 (0.62–1.54, P=0.84) in amplified tumours, and 1.36 (0.73–2.54, P=0.165) for the unadjusted interaction (Supplementary Table S9). The unmodified estimator behaves differently in two of the three strata: because the between-study variance is zero and the weighted residual mean square is below one, it rescales the variance downward and so narrows rather than widens the interval, to 0.98 (0.83–1.15, P=0.60) in amplified tumours and 1.36 (1.13–1.65, P=0.020) for the interaction. We report the modified form throughout because a variance estimate from three studies should not be allowed to make an interval narrower than the fixed-effect one. The non-amplified estimate also depends on one cohort: of the three, only GSE62564 reaches significance within the stratum (HR 0.67, P=0.007; GSE85047 P=0.087, TARGET-NBL P=0.423), and omitting it moves the pooled estimate to 0.83 (0.67–1.03, P=0.090). We therefore report the direction and consistency of the stratified estimates as the finding, and treat the effect modification by MYCN as suggestive rather than established.

Within-cohort interaction tests were individually underpowered (ratio of hazard ratios 1.39, 1.42 and 1.22; P=0.256, 0.078 and 0.519), but pooling them gave a consistent and statistically supported effect: 1.36 (1.03–1.81, P=0.032) unadjusted and 1.36 (1.01–1.82, P=0.044) after adjustment for age and stage, with I²=0% in both cases (Fig. 1C); no single cohort reaches nominal significance on its own (Supplementary Fig. S2E). Pooled Kaplan– Meier display shows the same pattern (non-amplified: n=732, 150 deaths, log-rank P=0.008; amplified: n=180, 106 deaths, P=0.521; Fig. 1D), as do the within-cohort median-split tests (Supplementary Fig. S2F).

The immune contrast associated with MYCN was broader than CLEC10A alone. Eleven of 12 prespecified genes were meta-estimable and all were lower in MYCN-amplified tumours (Supplementary Fig. S2A; TRAC was absent or invariant in two cohorts and is shown for TARGET-NBL only in Supplementary Fig. S2C). Random-effects module estimates were −0.86 (−1.13 to −0.59) for antigen presentation and −0.82 (−1.12 to −0.52) for the T-cell-inflamed module (Supplementary Fig. S2B), and both were lower in every cohort taken separately (Supplementary Fig. S2D). These are observational contrasts and do not establish direct MYCN-mediated suppression of any of these genes.

### CLEC10A detection in single cells is confounded by sequencing depth and cell-type composition

The myeloid/APC compartment of GSE147766 contained 8,219 cells from 17 biological patients, of which 8,092 cells from 14 patients were MYCN non-amplified. As expected, the patient-level fraction of CLEC10A-positive cells was highest in cDC2-like cells (median 78.1%) and near zero in cDC1-like cells (median 0%, four of the six evaluable patients having none; 0.4% of all cDC1-like cells), supporting the specificity of the marker (Fig. 2A–B).

**Figure 2.**
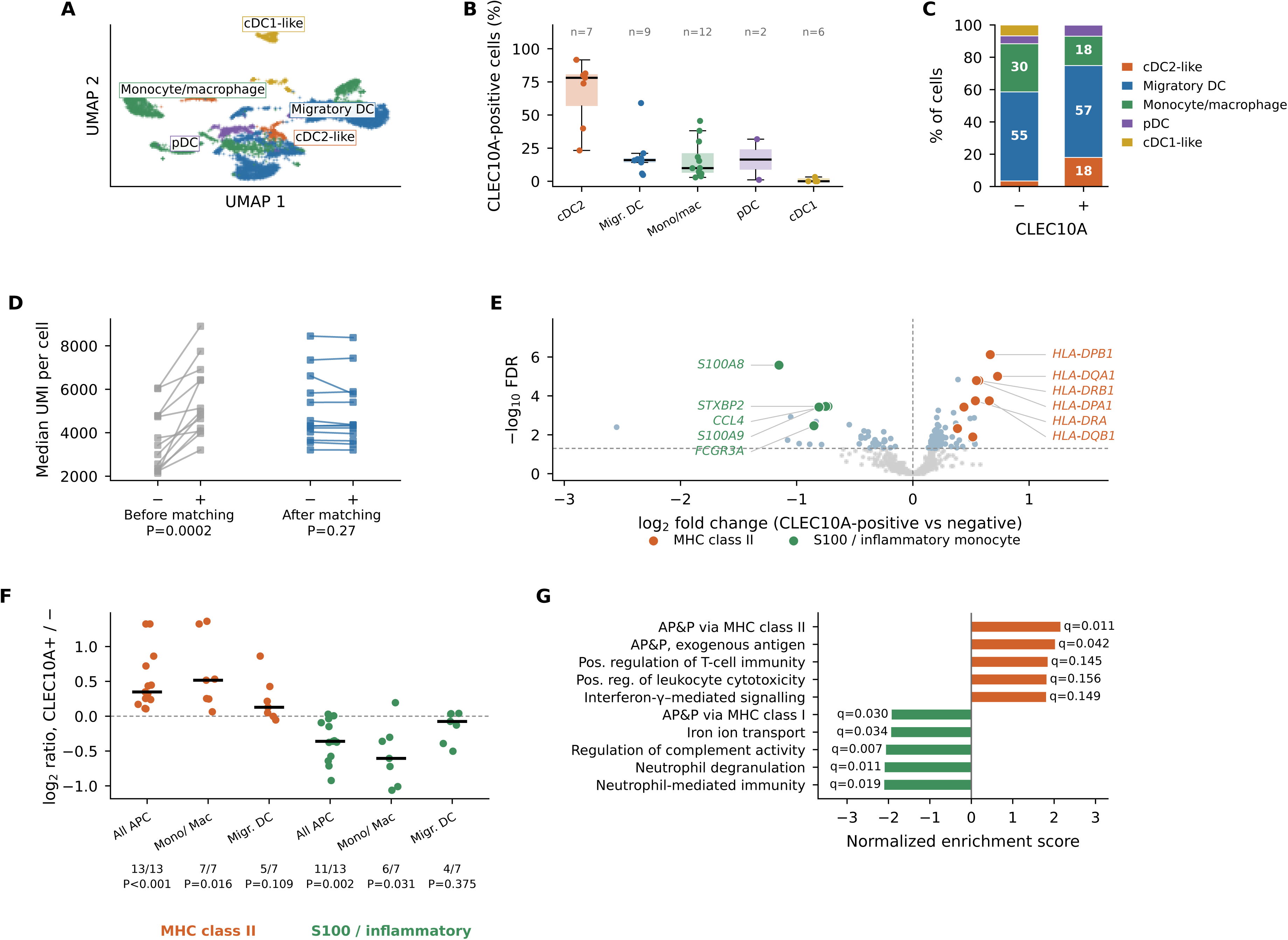
CLEC10A marks an MHC class II-high, alarmin-low state within tumour myeloid cells. MYCN non-amplified tumours; annotation was defined without CLEC10A. **A**, Myeloid/APC UMAP. **B**, Patient-level fraction of CLEC10A-positive cells by subtype, restricted to patients contributing ≥20 cells of that subtype; n is the number of such patients. **C**, Subtype composition of CLEC10A-negative and CLEC10A-positive cells; despite the highest detection rate in cDC2-like cells, only 18% of CLEC10A-positive cells are cDC2-like. **D**, Median UMI per cell by CLEC10A status, before and after matching; each line is one patient. **E**, Depth- and subtype-matched, biological-patient-paired pseudobulk differential expression (13 patient pairs, 1,216 matched cell pairs). CLEC10A is excluded because it defines the groups. Labelled genes are the most significant members of each highlighted set, placed in a column with leader lines to their points. **F**, Patient-level MHC class II and S100/inflammatory module scores, overall and within single subtypes; numbers below each group give patients concordant with the overall direction and the paired Wilcoxon P. **G**, Pre-ranked gene-set enrichment on the matched contrast after removal of ribosomal protein genes; GO term names are abbreviated and given in full in Supplementary Table S3. FDR, false-discovery rate; NES, normalised enrichment score.

Two features of the data make the conventional CLEC10A-positive versus negative contrast unsafe. First, because cDC2-like cells are a minority of the myeloid compartment, only 18% of CLEC10A-positive cells were cDC2-like; the largest contributors were migratory DC (57%) and monocytes/macrophages (18%), and the composition of the positive and negative pools differed substantially (Fig. 2C). A naive contrast therefore partly measures cell identity rather than cell state. Second, CLEC10A-positive cells were more deeply sequenced than CLEC10A-negative cells in every patient examined (median UMI ratio 1.48, paired Wilcoxon P=0.0002, 13/13 patients; Fig. 2D), and within-patient logistic regression gave an odds ratio for CLEC10A detection of 2.48 (2.28–2.70) per 1-SD higher log UMI (P=4×10⁻⁹⁶; Supplementary Fig. S3A).

Matching each CLEC10A-positive cell to a depth-matched CLEC10A-negative cell of the same subtype and the same patient produced 1,216 matched pairs across 14 patients, of which 13 patients and 1,214 pairs met the pre-set requirement of at least 10 matched cells per group (pairs by subtype in Supplementary Fig. S4C) and entered the differential expression, and removed the depth imbalance (cell-level median UMI 4,003 versus 4,037; patient-level paired Wilcoxon P=0.27). The consequences were substantial. The number of genes significant at FDR<0.05 fell from 395 to 107 (Supplementary Fig. S3B–C; full results in Supplementary Tables S5 and S1), and the strongest positive Hallmark signal of the unmatched analysis, MYC targets v1, collapsed from NES 2.14 (q<0.001) to NES 1.29 (q=0.83) (Supplementary Fig. S3D; Supplementary Table S4).

Matching alters four things at once — sequencing depth, subtype composition, the number of cells compared, and through that the genes surviving independent filtering — so we separated them, running the same patient-paired model on the same cells under five designs and scoring each identically (Methods; Supplementary Table S11). Reducing the negatives to the number of positives alone took the significant gene count from 395 to a median of 289 over five seeds; matching on subtype alone gave 207, on depth alone 157, and on both 113, close to the 107 obtained with the single seed used for the reported analysis (Supplementary Fig. S3E). The fall in gene count is therefore not attributable to any one of the three.

The MYC-target signal behaves differently, and its attribution is unambiguous. Scored as a competitive rank test of the gene set against all other tested genes, it was +6.9 in the unmatched contrast, +5.2 with cell number alone controlled, +3.7 with subtype alone and +2.0 with depth alone; adding subtype on top of depth changed nothing further (+2.3). Essentially all of it is attributable to sequencing depth, and we therefore describe the enrichment of MYC targets in CLEC10A-positive APCs as an artefact of library depth. The MHC class II signal was the one quantity invariant to every correction: +7.0 unmatched, and +7.1 to +7.2 under each of the four matched designs.

### After matching, CLEC10A marks an MHC class II-high, alarmin-low myeloid state

The signal that survived matching was coherent. Of the 106 differentially expressed genes other than CLEC10A itself, 72 were higher in CLEC10A-positive cells and 34 lower, and seven of the eight most significant upregulated genes were MHC class II — HLA-DPB1, HLA-DQA1, HLA-DRB1, HLA-DPA1, HLA-DRA, HLA-DQB1 and HLA-DMA — the eighth being HCST. The two remaining class II genes in the tested set, CD74 and HLA-DRB5, were also significant, ranked 18th and 33rd of the 72. The genes lower in CLEC10A-positive cells were dominated by the S100 alarmin and inflammatory monocyte programme: S100A8, STXBP2, CCL4, S100A9 and FCGR3A were all significant and are the first, second, third, fourth and eighth most significant of the 34. Two further members of that programme moved in the same direction without reaching significance — SERPINA1 (log₂ −0.40, q=0.07) and S100A12 (log₂ −1.14), the latter not passing independent filtering (Fig. 2E).

The MHC class II module score was higher in CLEC10A-positive cells in 13 of 13 patients (median log₂ ratio 0.35, paired Wilcoxon P=0.0002), and the S100 module was lower in 11 of 13 (median log₂ ratio −0.36, P=0.002). Critically, both effects were reproduced within monocytes/macrophages alone — 7 of 7 patients for MHC class II (log₂ 0.52, P=0.016) and 6 of 7 for S100 (log₂ −0.61, P=0.031) — establishing that the state is intrinsic to a single myeloid cell type and cannot be explained by cDC2 admixture. In migratory DC the same directions were seen but did not reach significance (5 of 7, P=0.109; 4 of 7, P=0.375; Fig. 2F, Supplementary Fig. S4A).

Gene-set enrichment on the matched contrast, after removal of ribosomal protein genes, ranked MHC class II antigen processing and presentation first among positively enriched terms (NES 2.16, q=0.011), followed by presentation of exogenous antigen (NES 2.03, q=0.042), positive regulation of T-cell-mediated immunity (NES 1.86, q=0.145), positive regulation of leukocyte-mediated cytotoxicity (NES 1.83, q=0.156) and interferon-γ-mediated signalling (NES 1.82, q=0.149). Negatively enriched terms were neutrophil degranulation (NES −2.10, q=0.011), neutrophil-mediated immunity (NES −2.11, q=0.019), regulation of complement activation (NES −2.07, q=0.007) and iron ion transport (NES −1.95, q=0.034). Notably, antigen processing and presentation via MHC class I ran in the opposite direction to class II (NES −1.93, q=0.030), indicating that CLEC10A-positive cells are not globally antigen-presentation-high but specifically class II-high (Fig. 2G). Omitting each patient in turn preserved the direction of every one of the 106 significant non-CLEC10A genes (sign concordance 1.00 in all 13 leave-one-patient-out fits; Supplementary Fig. S4B).

Repeating the identical pipeline on all patients rather than the MYCN non-amplified subset gave the same result (Supplementary Fig. S5; full results in Supplementary Table S2). That scope yielded 1,232 matched pairs across 15 patients, of which 14 patients and 1,230 pairs met the same minimum of 10 matched cells per group, 120 differentially expressed genes excluding CLEC10A, and the same MHC class II-dominated top set; log₂ fold changes correlated across the two scopes at r=0.986 and all 106 genes significant in the primary analysis retained their direction. Gene-set enrichment was also preserved, with MHC class II antigen processing and presentation the strongest term (NES 2.36, q<0.001) and translation-related terms remaining weak (q≥0.08), confirming that the depth correction holds in the wider scope.

### In tissue, CLEC10A-positive myeloid bins occupy T-cell-richer neighbourhoods, and this survives depth matching

Having established what CLEC10A marks within a cell, we asked where those cells sit. Six Visium HD sections from five patients met the prespecified immune-content criteria, comprising 53,433 to 166,127 in-tissue bins each at 8 µm resolution (Supplementary Table S7); one section is shown in Fig. 3A.

**Figure 3.**
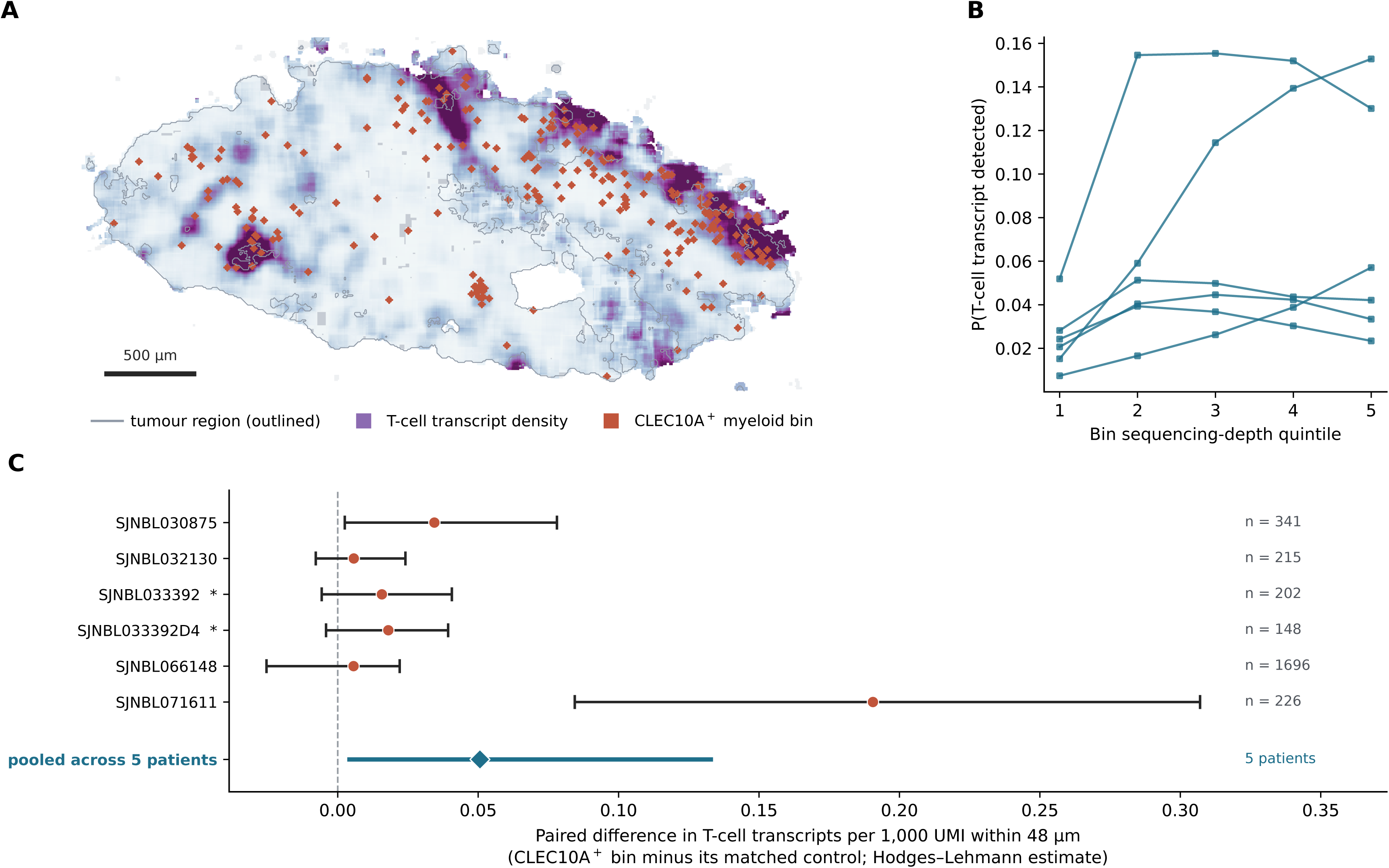
CLEC10A-positive myeloid bins occupy T-cell-richer neighbourhoods. Visium HD, 8 µm bins, six neuroblastoma sections from five patients (GSE332618). **A**, One section (SJNBL071611, 76,964 in-tissue bins): the tumour region is outlined, T-cell transcript density is shown in purple and CLEC10A-positive myeloid bins in orange, the latter dilated to 24 µm for visibility. The field shows the main tissue body, which contains 92% of in-tissue bins; detached fragments lie outside it and are not displayed, although all in-tissue bins enter the analysis. Scale bar, 500 µm. **B**, Probability that a bin is called T-cell positive, by bin sequencing-depth quintile; one line per section. Detection follows depth in five of the six sections, so any detection-based spatial outcome is depth-driven; the distance-based alternative and its failure are shown in Supplementary Figure S6B–D. **C**, Paired difference in the depth-normalised outcome — T-cell transcripts per 1,000 UMI within 48 µm, excluding the bin itself — between each CLEC10A-positive myeloid bin and the CLEC10A-negative myeloid bin matched to it on myeloid count and log total UMI within section. Points are Hodges–Lehmann estimates and bars are 95% intervals from a spatial block bootstrap over 200 µm tiles; n is the number of matched pairs. Asterisks mark the two sections contributed by one patient. The pooled estimate (diamond) is from a hierarchical bootstrap that resamples the five patients and then tiles within each, and is the quantity on which inference rests; per-section intervals are shown for completeness, are listed in Supplementary Table S10b, and are not independent of one another.

The same depth confound operates on the spatial data. The probability that an 8 µm bin was called T-cell positive rose with bin sequencing depth in five of the six sections, the ratio between the highest and the lowest depth quintile ranging from 0.97 to 10.1 (Fig. 3B), and bin depth was spatially autocorrelated, predicting the mean depth of the surrounding 48 µm ring at Spearman ρ 0.783–0.891 and of a 24 µm ring at 0.871–0.942 (Supplementary Fig. S6A). Outcomes built on T-cell detection are therefore partly depth-driven, and the natural choice — distance to the nearest T-cell-positive bin — is on an 8 µm lattice additionally confined to a small set of discrete values (8.0, 11.3, 16.0, 17.9, 22.6 µm; Supplementary Fig. S6B), so group medians can fall only on those values and coincided in four of the six sections after matching (Supplementary Fig. S6D). We therefore used a depth-normalised outcome: T-cell transcripts per 1,000 UMI within 48 µm of each bin, excluding the bin itself. CLEC10A positivity is itself a detection call, so each CLEC10A-positive myeloid bin was matched within section to a CLEC10A-negative myeloid bin on myeloid count and log total UMI; matching equalised the depth of the neighbourhood as well as of the bin (ratio of mean neighbourhood UMI 0.963–1.003, Supplementary Fig. S6E). The unmatched comparisons are reported alongside the matched ones throughout (Supplementary Fig. S6C,F).

Neighbourhoods of 48 µm overlap — 70% to 100% of analysed bins per section lie within 96 µm, twice the neighbourhood radius, of another analysed bin — and two of the six sections come from one patient. Inference is therefore at the level of the patient, with intervals from a spatial block bootstrap over contiguous tiles nested inside a bootstrap over patients (Methods).

The paired difference was positive in all six sections and in all five patients. Per section, the Hodges–Lehmann estimate of the paired difference and its block-bootstrap interval were 0.034 (0.003 to 0.078) for SJNBL030875, 0.006 (−0.008 to 0.024) for SJNBL032130, 0.016 (−0.006 to 0.041) for SJNBL033392, 0.018 (−0.004 to 0.039) for SJNBL033392D4, 0.006 (−0.025 to 0.022) for SJNBL066148 and 0.191 (0.084 to 0.307) for SJNBL071611 T-cell transcripts per 1,000 UMI (Fig. 3C). Pooled over the five patients, the difference was 0.051 (95% bootstrap interval 0.003 to 0.134), and the estimate was stable to the choice of tile size between 96 µm and 400 µm. Ignoring the spatial dependence would have made these intervals 1.3 to 2.2 times narrower than they are.

The pooled interval excludes zero, but only narrowly and only with all five patients present. The effect is dominated by SJNBL071611, and omitting any one of three patients brings zero inside the interval (Supplementary Table S10). What the data support is a consistent direction across five patients, not a reliably estimated magnitude.

Matching equalised not only each bin’s own depth but the depth of its neighbourhood, the ratio of mean neighbourhood UMI between CLEC10A-positive bins and their matched controls lying between 0.963 and 1.003 across sections, so the groups are balanced on neighbourhood depth as well as on their own. Balance of the mean does not establish balance of the distribution, and residual confounding through local composition is not excluded. The one section without a difference, SJNBL066148, is also the section in which myeloid-positive bins make up 59.6% of tissue and CLEC10A-positive bins are an order of magnitude more numerous than elsewhere (1,697 versus 149–343); where myeloid cells are distributed throughout the tissue there is little positional contrast left to detect.

These data place the CLEC10A-associated myeloid state in T-cell-richer tissue, measured over a fixed physical radius and with the depth confound addressed. They do not show that these myeloid cells prime, activate or recruit T cells; co-location at 8 µm is compatible with any of those and with none of them.

## Discussion

CLEC10A has been read in neuroblastoma, as elsewhere, as a count of cDC2 [14,19,20]. These data indicate that in tumour tissue it is better read as a marker of a state. Once the two artefacts that dominate marker-positive versus marker-negative comparisons are removed, what CLEC10A identifies is an MHC class II-high, alarmin-low transcriptional programme that is present across myeloid cell types rather than confined to one lineage, that occupies T-cell-richer tissue, and whose association with survival appears only where *MYCN* is not amplified.

The state is not a property of which broad cell types are present. CLEC10A-positive myeloid cells are MHC class II-high and S100 alarmin-low, and both directions reproduce within monocytes and macrophages considered alone — class II in 7 of 7 patients, S100 in 6 of 7 — so the signal cannot be explained by cDC2 admixture, and indeed only 18% of CLEC10A-positive cells were cDC2-like. Monocytes and macrophages are themselves heterogeneous [22], so this does not exclude every compositional explanation; it excludes the one the marker invites. The accompanying reductions in complement and neutrophil-like inflammatory programmes argue for a specific phenotype rather than generic activation. Being alarmin-low is itself informative, because S100A8, S100A9 and S100A12 define the inflammatory monocyte programme that characterises suppressive myeloid states [39,40]; the CLEC10A-associated state runs against that programme rather than with it. The module we scored is broader than the three alarmins, so this is a statement about the programme as a whole.

That this is a class II programme carries more weight in neuroblastoma than it would in most tumours. Neuroblastoma presents poorly through MHC class I. Its somatic mutation burden is among the lowest of any cancer, so few neoantigens are available [7], and class I expression on tumour cells is low, a defect tied to *MYCN* amplification in work from 1986 [8]. Where class I presentation is constrained, the class II–CD4 axis is what remains, and MHC class II-restricted help can by itself drive protective antitumour immunity [17]. A readout of intratumoural class II capacity is therefore of more than descriptive interest here. One feature of the state points the same way and was not anticipated: antigen processing and presentation via MHC class I ran in the *opposite* direction to class II within the same cells. Whether that reflects a division of labour inside the myeloid compartment, or the same pressures that lower class I on neuroblastoma cells acting on myeloid cells as well, cannot be settled from one dataset; we offer it as a hypothesis for replication rather than as a finding.

In tissue, this state is found where T cells are. At 8 µm resolution, CLEC10A-positive myeloid bins occupy neighbourhoods richer in T-cell transcripts than myeloid bins matched for sequencing depth and myeloid content, in all six sections and all five patients. The direction is the robust part. The magnitude is not: one patient contributes most of it, the pooled interval excludes zero only when all five are present, and with five patients a bootstrap interval is itself a rough instrument. The claim is correspondingly narrow — the state is positioned near T cells rather than away from them. Proximity within a 48 µm radius is not contact, a bin is not a cell, and nothing here shows that anything passes between them.

The survival association behaves in a way that fits this reading, and in doing so resolves an apparent inconsistency. Taken cohort by cohort CLEC10A looks unreliable: the association is clear in GSE85047 and in GSE62564 and absent in TARGET-NBL. That is a composition effect. TARGET-NBL is heavily enriched for high-risk and stage 4 disease (Table 1), and once the three cohorts are stratified by *MYCN* status the between-cohort heterogeneity is no longer detectable (I² from 82% to 0%) and the estimates align in direction. How strongly that can be asserted is limited by having three cohorts: the between-study variance is estimated as zero, so the random-effects interval is really a fixed-effect interval, and an estimator that prices in the uncertainty of that variance widens the stratum estimate to 0.77 (0.53–1.12) and the interaction to 1.36 (0.73–2.54). Within the non-amplified stratum only GSE62564 is individually significant. The consistent direction across three cohorts and two strata is the substance of the result; the effect modification itself is suggestive, and we do not claim it as established. It remains true that CLEC10A is not independently prognostic after clinical adjustment, and we do not propose it as a stand-alone biomarker. Its potential use is different in kind: a readout, available in the bulk datasets that most neuroblastoma cohorts consist of, of an immune property that single-cell work can otherwise measure in only a handful of patients.

This reading converges with, rather than contradicts, the annotation adopted in the harmonised neuroblastoma single-cell reference atlas [14]. That atlas calls cDC2 by CD1C, CD1E and CLEC10A, which is the conventional use of the marker, but it also places a separate CLEC10A-expressing population inside the monocyte compartment, annotated as monocytes in transition to macrophages. This is not independent replication: the dataset re-analysed here [15] is one of the seven studies NBAtlas integrates, and with 19 of its 68 samples it is the largest single contributor. What it does show is that the assignment survives a different annotation pipeline applied to a corpus six times larger, rather than being a product of how we annotated these cells. What a reference atlas is not built to establish — its purpose being to assign identity rather than to contrast states within one — is what CLEC10A expression means functionally inside those cells; the atlas does not link CLEC10A to MHC class II at any point, attributing the highest HLA expression instead to a FOLR2-positive macrophage population of the kind associated with T-cell infiltration in other tumours [42]. One point in that annotation deserves comment, because it runs against our interpretation and would otherwise be left open. The atlas names IDO1 and LGALS2 alongside CLEC10A among the genes marking its CLEC10A-expressing monocyte cluster [14], and IDO1 is immunoregulatory [43]; if CLEC10A tracked IDO1 at the single-cell level, a class II-high, T-cell-supportive reading would be hard to sustain. In these data it does not. IDO1 was not higher in CLEC10A-positive cells even in the naive contrast, in which the depth confound biases genes upward (log₂ fold change −0.71, P=0.30), and was too sparsely detected to test after matching; LGALS2 was nominally higher before matching (log₂ 0.73, P=0.009) but not after it (log₂ 0.58, P=0.12) and was flat within monocytes and macrophages alone (log₂ 0.02, P=0.96). Of the markers the atlas assigns to that cluster, only the MHC class II transcripts it does *not* assign to CLEC10A survive depth matching.

These observations form a single axis. The prespecified antigen-presentation module is lower in *MYCN*-amplified tumours; CLEC10A marks an MHC class II-high myeloid state at the single-cell level; that state sits where T cells are; and the survival association attached to CLEC10A is present only where the immune context is preserved — in *MYCN* non-amplified disease. The parsimonious interpretation is that CLEC10A is a readout of intratumoural class II antigen-presentation capacity, and that this capacity carries prognostic information only when *MYCN* amplification has not already dictated the outcome.

What this chain does not contain is the step that would make it mechanistic. We have a prognostic association stratified by *MYCN*, a cell state defined at single-cell resolution with its two main artefacts removed, and a spatial relationship between that state and T cells. We do not have evidence that these myeloid cells present antigen to, activate, or expand T cells. That step cannot be supplied by any of the public data used here: co-location is not interaction, transcript is not protein, and an association across tumours is not a demonstration of causation within one. It requires functional work, and we regard it as the central open question rather than a detail to be filled in later. CLEC10A is also not an inert marker. Its ligand is terminal N-acetylgalactosamine, including the tumour-associated Tn antigen [18], and engagement of CD45 on T cells by the receptor has been shown to attenuate effector T-cell responses [44]. A receptor that marks antigen-presentation capacity while also being able to restrain the T cells it meets is not a contradiction, but it does mean that the sign of any functional effect cannot be read off expression. Our claim concerns what CLEC10A expression *marks* on myeloid cells, not what ligation of the receptor *does*, and the two should not be conflated.

One methodological point bears statement, because it determines whether results of this kind can be believed. Grouping cells by detection of a marker makes sequencing depth a determinant of the *exposure* rather than of the outcome: here the odds of being called CLEC10A-positive rose 2.5-fold per standard deviation of log depth, and because the marker is enriched in one subtype the two pools differed in composition as well. Covariate adjustment of the kind MAST performs [30] cannot repair a contrast already formed on a depth-dependent split. The consequences were not subtle — 395 significant genes and a strong MYC-target signature before matching, 107 genes and no MYC-target enrichment after. Running the same model under designs that control depth, composition and cell number separately shows that the MYC-target signal tracks depth alone, that the gene-count drop reflects all three, and that the MHC class II signal moves under none of them; attribution of this kind is worth doing, because matching changes several things at once and the naive reading is that it changed only the one the analyst had in mind. The same mechanism operated on the spatial data, where a detection-based proximity outcome was depth-driven until it was replaced by one normalised to local depth. Because spatial and single-cell data are increasingly used to make proximity claims about marker-defined populations [41,42], we report the naive and corrected contrasts side by side throughout and would encourage the same practice. The survival analysis makes a parallel point: where cohorts differ in clinical composition, stratified meta-analysis is more informative than counting significant and non-significant cohorts.

Several limitations apply. All analyses are retrospective and observational; the cohorts differ in platform and in the covariates available, and adjustment attenuates the survival association (Supplementary Fig. S1E). The proportional-hazards assumption is violated for the CLEC10A term in GSE62564 and, univariably, in TARGET-NBL (Supplementary Fig. S1F), so the hazard ratios we report should be read as time-averaged effects rather than constant ones; because almost all deaths occur within five years the estimates are nevertheless stable across follow-up horizons (Supplementary Fig. S1G). We show that *MYCN* status modifies the CLEC10A–survival relationship, but not that *MYCN* suppresses CLEC10A or class II presentation directly. The single-cell analysis rests on 13 patient pairs from one atlas, *MYCN*-amplified tumours contributed too few APCs for a paired contrast, and a cDC2-restricted analysis was underpowered (4 eligible patients). One sample, NB37, is classified as *MYCN* non-amplified by the source study’s annotation, which we followed, but as amplified in the reference atlas, which assigns status by inference; it contributes 318 of the 8,092 cells in the scope and only 2 matched pairs, so it is excluded from the differential expression by the minimum-cell rule and cannot affect that result, but the scope statement depends on which annotation is taken. Matching necessarily discards unmatched cells and can only balance the confounders that are measured; residual differences in RNA content or capture efficiency may persist.

The spatial analysis carries its own limits. It rests on six sections from five patients, one patient contributing two; *MYCN* status is not recorded in the source annotation, so the spatial result could not be stratified by the variable that organises the rest of this study; an 8 µm bin contains one to a few cells, so a bin is not a cell and adjacency is not contact; the Visium HD probe panel covers 18,132 genes rather than the whole transcriptome; the pooled interval excludes zero only with all five patients present and is dominated by one of them, and a bootstrap interval built on five units is a rough instrument; and the dataset had no associated publication at the time of analysis, so it is cited by accession only. Finally, transcript detection is not protein abundance, and none of these data demonstrate function.

The obvious next steps follow from those limits. Flow or mass cytometry should test whether CLEC10A-positive tumour myeloid cells carry higher surface HLA-DR protein and lower S100A8/A9, in *MYCN*-amplified as well as non-amplified tumours. Because our claim is that CLEC10A *marks* a functional state rather than that it *creates* one, the decisive experiment is to sort CLEC10A-positive and CLEC10A-negative myeloid cells and compare their capacity to activate and polarise naive T cells, rather than to force CLEC10A expression, which tests a different proposition. Spatial data at single-cell resolution, and paired T-cell receptor repertoires, would establish whether co-location corresponds to clonal engagement. Until then, CLEC10A is best used as a contextual readout of myeloid class II antigen-presentation capacity in *MYCN* non-amplified neuroblastoma.

## Data Availability

All datasets analysed are publicly available: GSE85047, GSE62564, GSE147766 and GSE332618 from the Gene Expression Omnibus, and TARGET-NBL from the NCI TARGET data portal. Analysis code, figure source data, matched-cell and matched-bin assignments, gene-set definitions and quality-control outputs are deposited at Zenodo (doi:10.5281/zenodo.23686189).

https://doi.org/10.5281/zenodo.23686189

## Declarations

## Ethics approval and consent to participate

This study analysed only publicly available, de-identified human data from GEO (GSE85047, GSE62564, GSE147766, GSE332618) and the NCI TARGET programme. No new human material was collected and institutional review board approval was not required.

## Consent for publication

Not applicable.

## Availability of data and materials

All datasets analysed are publicly available: GSE85047, GSE62564, GSE147766 and GSE332618 from the Gene Expression Omnibus, and TARGET-NBL from the NCI TARGET data portal. GSE332618 had no associated publication at the time of analysis and is cited by accession. Analysis code, figure source data, matched-cell and matched-bin assignments, gene-set definitions and quality-control outputs are deposited at Zenodo (doi:10.5281/zenodo.23686189). The deposit also contains the superseded, pre-matching single-cell outputs and the pre-matching spatial outputs, so that the effect of the depth and composition corrections described in Supplementary Figures S3 and S6 can be inspected directly.

## Competing interests

The authors declare no competing interests.

## Funding

This research received no specific grant from any funding agency in the public, commercial or not-for-profit sectors.

## Authors’ contributions

S.T. conceived the study, assembled the datasets, performed all analyses and wrote the manuscript. K.Y., T.T., H.K., A.N., Y.K., S.W., T.Y., E.O. and M.I. reviewed the analyses and their interpretation and provided critical comment on the content of the manuscript. All authors read and approved the final version and agree to be accountable for all aspects of the work.

## Acknowledgements

We thank the investigators of GSE85047, GSE62564, GSE147766, GSE332618 and the NCI TARGET programme for making their data publicly available.

## Notes

### Competing Interest Statement

The authors have declared no competing interest.

### Author Declarations

The study used ONLY openly available human data, all of which were in the public domain before this work began. No application, request, screening or registration was required for access. Bulk tumour expression and outcome data: - Gene Expression Omnibus GSE85047 (https://www.ncbi.nlm.nih.gov/geo/query/acc.cgi?acc=GSE85047) - Gene Expression Omnibus GSE62564 (https://www.ncbi.nlm.nih.gov/geo/query/acc.cgi?acc=GSE62564) - NCI Therapeutically Applicable Research to Generate Effective Treatments (TARGET) neuroblastoma programme, open-access tier (https://www.cancer.gov/ccg/research/genome-sequencing/target) Single-cell RNA sequencing data: - Gene Expression Omnibus GSE147766 (https://www.ncbi.nlm.nih.gov/geo/query/acc.cgi?acc=GSE147766) Visium HD spatial transcriptomic data: - Gene Expression Omnibus GSE332618 (https://www.ncbi.nlm.nih.gov/geo/query/acc.cgi?acc=GSE332618) All datasets are de-identified and openly downloadable. No new human data, samples or participants were involved, and no institutional review board approval was therefore required.

