## Supplementary Figures S1-S6 and Supplementary Tables S1-S11 for "CLEC10A marks an MHC class II-high tumour myeloid state that lies in T-cell-rich neighbourhoods, and its survival association is seen in MYCN non-amplified neuroblastoma": Supplementary_Figure_S1.pdf

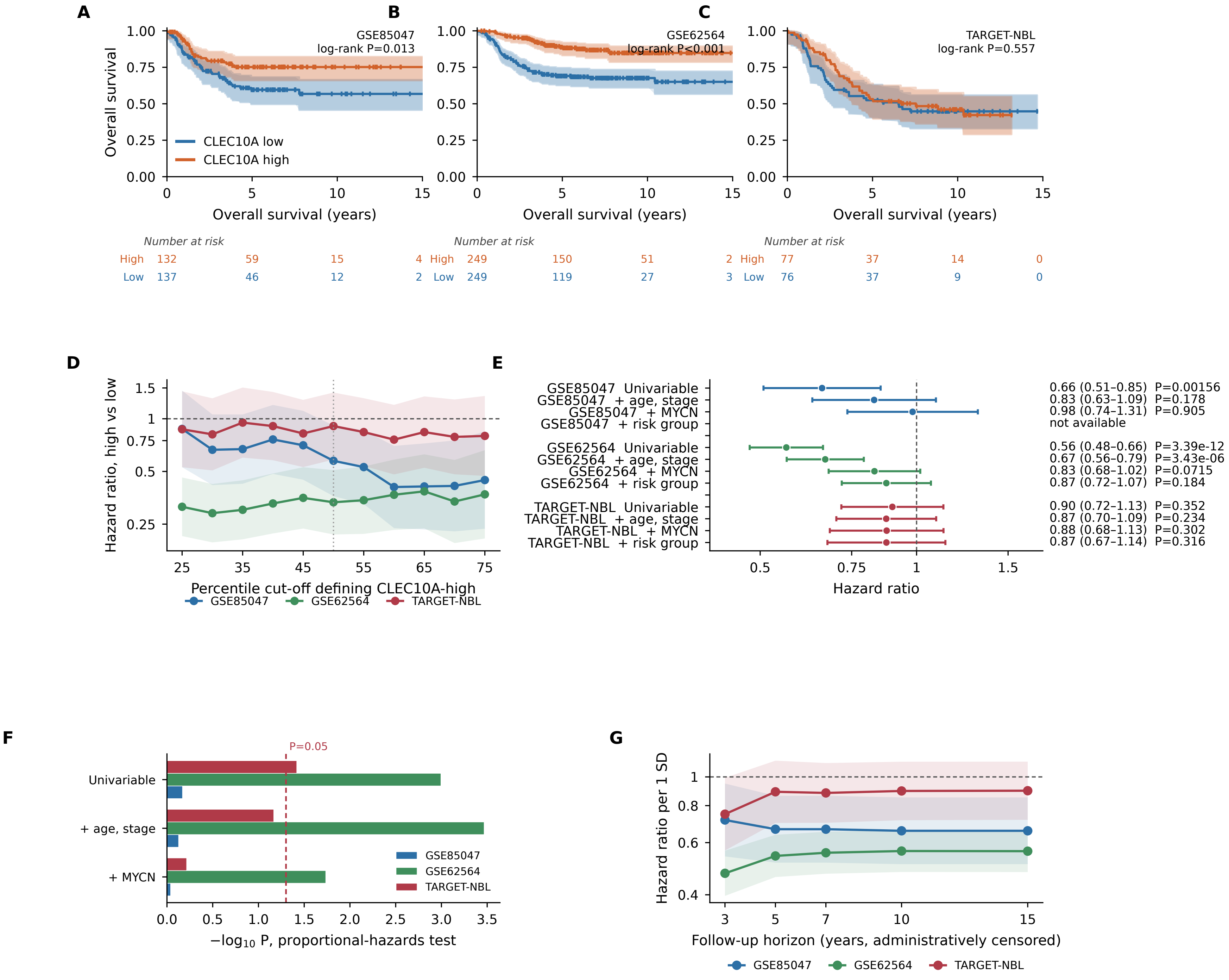
A
B
C
GSE85047
log-rank P=0.013
GSE62564
log-rank P<0.001
TARGET-NBL
log-rank P=0.557
Overall survival
CLEC10A low
CLEC10A high
Overall survival (years)
Number at risk
High 132 59 15
Low 137 46 12
4 High 249 150 51
2 Low 249 119 27
2 High 77 37 14 0
3 Low 76 37 9 0
D
Hazard ratio, high vs low
Percentile cut-off defining CLEC10A-high
GSE85047
GSE62564
TARGET-NBL
E
GSE85047 Univariable
GSE85047 + age, stage
GSE85047 + MYCN
GSE85047 + risk group
GSE62564 Univariable
GSE62564 + age, stage
GSE62564 + MYCN
GSE62564 + risk group
TARGET-NBL Univariable
TARGET-NBL + age, stage
TARGET-NBL + MYCN
TARGET-NBL + risk group
0.66 (0.51–0.85) P=0.00156
0.83 (0.63–1.09) P=0.178
0.98 (0.74–1.31) P=0.905
not available
0.56 (0.48–0.66) P=3.39e-12
0.67 (0.56–0.79) P=3.43e-06
0.83 (0.68–1.02) P=0.0715
0.87 (0.72–1.07) P=0.184
0.90 (0.72–1.13) P=0.352
0.87 (0.70–1.09) P=0.234
0.88 (0.68–1.13) P=0.302
0.87 (0.67–1.14) P=0.316
Hazard ratio
F
P=0.05
Univariable
+ age, stage
+ MYCN
GSE85047
GSE62564
TARGET-NBL
−log10 P, proportional-hazards test
G
Hazard ratio per 1 SD
Follow-up horizon (years, administratively censored)
GSE85047
GSE62564
TARGET-NBL
