## Supplementary Figures S1-S6 and Supplementary Tables S1-S11 for "CLEC10A marks an MHC class II-high tumour myeloid state that lies in T-cell-rich neighbourhoods, and its survival association is seen in MYCN non-amplified neuroblastoma": Supplementary_Figure_S2.pdf

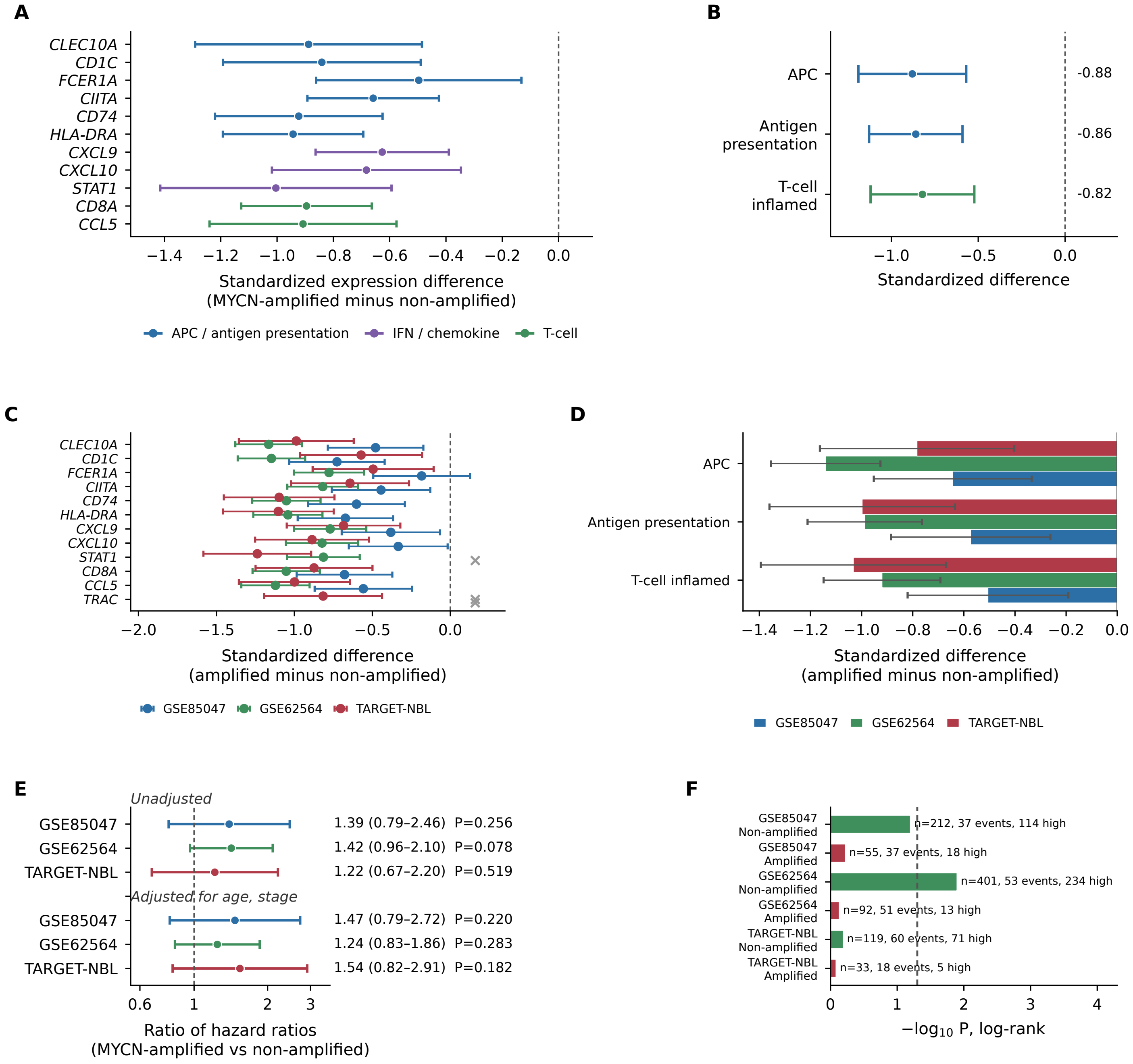
A
CLEC10A
CD1C
FCER1A
CIITA
CD74
HLA-DRA
CXCL9
CXCL10
STAT1
CD8A
CCL5
−1.4 −1.2 −1.0 −0.8 −0.6 −0.4 −0.2 0.0
Standardized expression difference
(MYCN-amplified minus non-amplified)
APC / antigen presentation
IFN / chemokine
T-cell
B
APC
Antigen presentation
T-cell inflamed
-0.88
-0.86
-0.82
−1.0 −0.5 0.0
Standardized difference
C
CLEC10A
CD1C
FCER1A
CIITA
CD74
HLA-DRA
CXCL9
CXCL10
STAT1
CD8A
CCL5
TRAC
−2.0 −1.5 −1.0 −0.5 0.0
Standardized difference
(amplified minus non-amplified)
GSE85047
GSE62564
TARGET-NBL
D
APC
Antigen presentation
T-cell inflamed
−1.4 −1.2 −1.0 −0.8 −0.6 −0.4 −0.2 0.0
Standardized difference
(amplified minus non-amplified)
GSE85047
GSE62564
TARGET-NBL
E
Unadjusted
GSE85047
GSE62564
TARGET-NBL
1.39 (0.79–2.46) P=0.256
1.42 (0.96–2.10) P=0.078
1.22 (0.67–2.20) P=0.519
Adjusted for age, stage
GSE85047
GSE62564
TARGET-NBL
1.47 (0.79–2.72) P=0.220
1.24 (0.83–1.86) P=0.283
1.54 (0.82–2.91) P=0.182
0.6 1 2 3
Ratio of hazard ratios
(MYCN-amplified vs non-amplified)
F
GSE85047 Non-amplified
n=212, 37 events, 114 high
GSE85047 Amplified
n=55, 37 events, 18 high
GSE62564 Non-amplified
n=401, 53 events, 234 high
GSE62564 Amplified
n=92, 51 events, 13 high
TARGET-NBL Non-amplified
n=119, 60 events, 71 high
TARGET-NBL Amplified
n=33, 18 events, 5 high
0 1 2 3 4
−log10 P, log-rank
