## Supplementary Figures S1-S6 and Supplementary Tables S1-S11 for "CLEC10A marks an MHC class II-high tumour myeloid state that lies in T-cell-rich neighbourhoods, and its survival association is seen in MYCN non-amplified neuroblastoma": Supplementary_Figure_S3.pdf

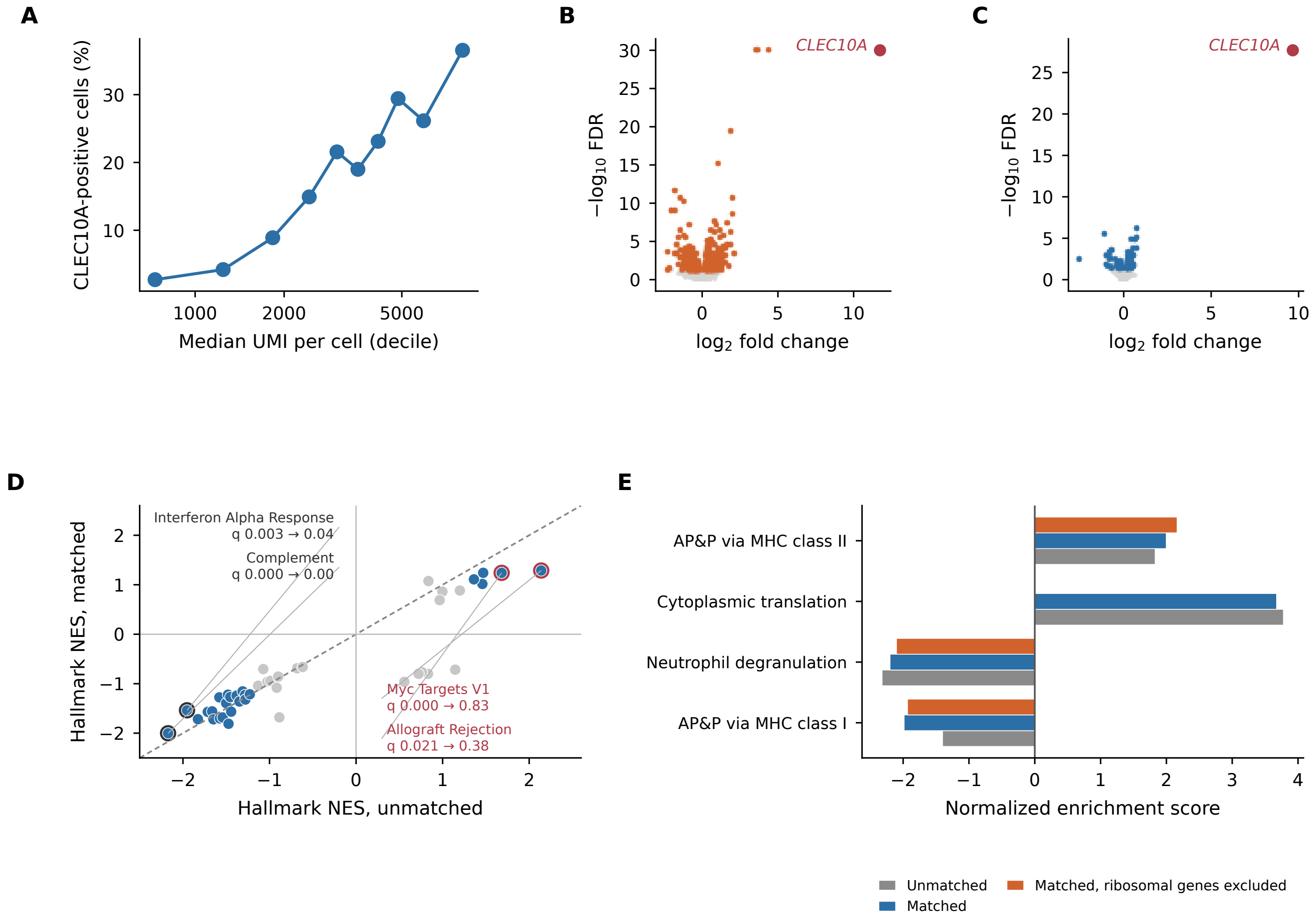
A
CLEC10A-positive cells (%)
Median UMI per cell (decile)
B
CLEC10A
−log10 FDR
log2 fold change
C
CLEC10A
−log10 FDR
log2 fold change
D
Interferon Alpha Response
q 0.003 → 0.04
Complement
q 0.000 → 0.00
Myc Targets V1
q 0.000 → 0.83
Allograft Rejection
q 0.021 → 0.38
Hallmark NES, matched
Hallmark NES, unmatched
E
AP&P via MHC class II
Cytoplasmic translation
Neutrophil degranulation
AP&P via MHC class I
Normalized enrichment score
Unmatched
Matched
Matched, ribosomal genes excluded
