## Supplementary Figures S1-S6 and Supplementary Tables S1-S11 for "CLEC10A marks an MHC class II-high tumour myeloid state that lies in T-cell-rich neighbourhoods, and its survival association is seen in MYCN non-amplified neuroblastoma": Supplementary_Figure_S4.pdf

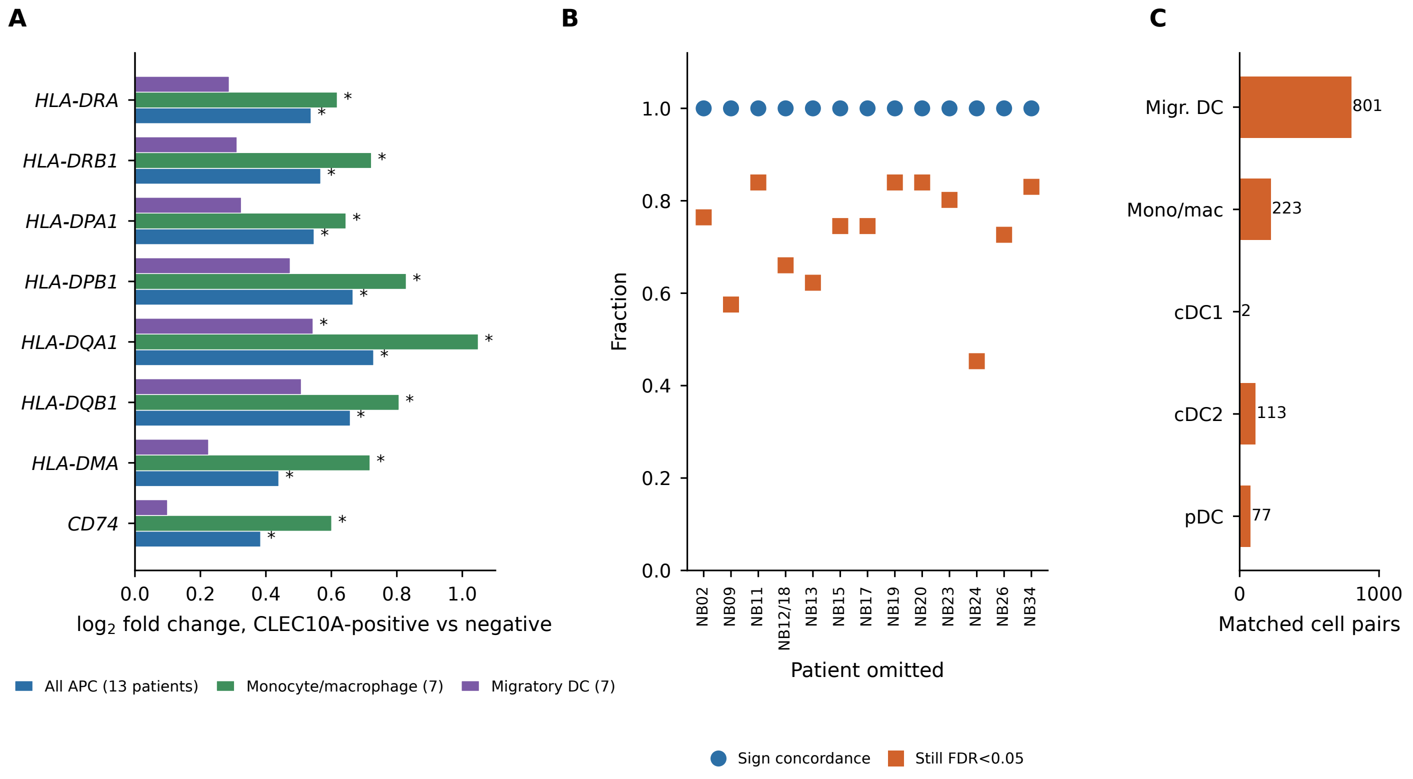
A
HLA-DRA
HLA-DRB1
HLA-DPA1
HLA-DPB1
HLA-DQA1
HLA-DQB1
HLA-DMA
CD74
0.0
0.2
0.4
0.6
0.8
1.0
log2 fold change, CLEC10A-positive vs negative
All APC (13 patients)
Monocyte/macrophage (7)
Migratory DC (7)
B
Fraction
1.0
0.8
0.6
0.4
0.2
0.0
NB02
NB09
NB11
NB12/18
NB13
NB15
NB17
NB19
NB20
NB23
NB24
NB26
NB34
Patient omitted
Sign concordance
Still FDR<0.05
C
Migr. DC
Mono/mac
cDC1
cDC2
pDC
801
223
2
113
77
0
1000
Matched cell pairs
