## Supplementary Figures S1-S6 and Supplementary Tables S1-S11 for "CLEC10A marks an MHC class II-high tumour myeloid state that lies in T-cell-rich neighbourhoods, and its survival association is seen in MYCN non-amplified neuroblastoma": Supplementary_Figure_S5.pdf

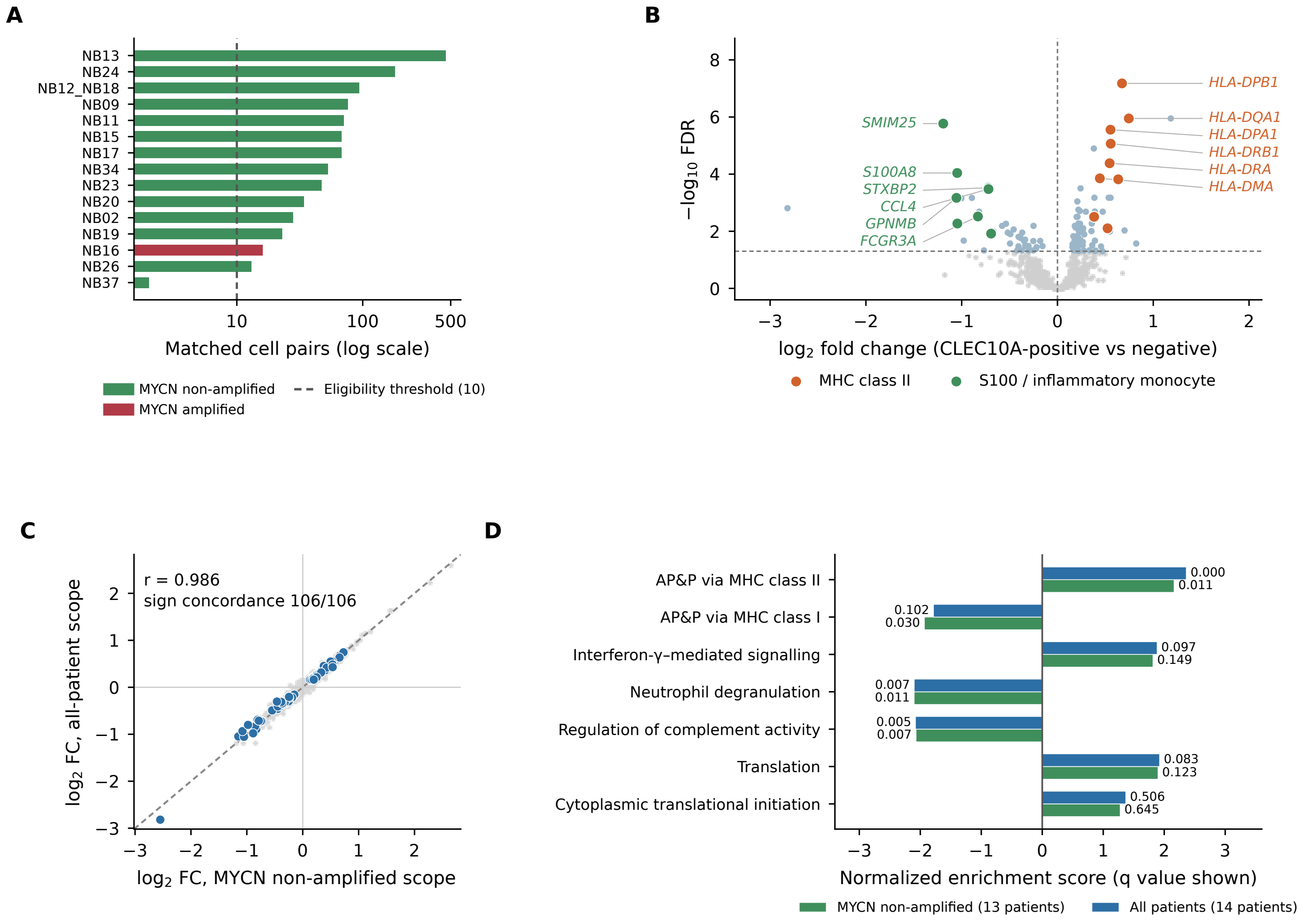

A
NB13
NB24
NB12_NB18
NB09
NB11
NB15
NB17
NB34
NB23
NB20
NB02
NB19
NB16
NB26
NB37
10
100
500
Matched cell pairs (log scale)
MYCN non-amplified
MYCN amplified
Eligibility threshold (10)
B
−log10 FDR
8
6
4
2
0
HLA-DPB1
HLA-DQA1
HLA-DPA1
HLA-DRB1
HLA-DRA
HLA-DMA
SMIM25
S100A8
STXBP2
CCL4
GPNMB
FCGR3A
−3
−2
−1
0
1
2
log2 fold change (CLEC10A-positive vs negative)
MHC class II
S100 / inflammatory monocyte
C
r = 0.986
sign concordance 106/106
log2 FC, all-patient scope
log2 FC, MYCN non-amplified scope
D
AP&P via MHC class II
AP&P via MHC class I
Interferon-γ–mediated signalling
Neutrophil degranulation
Regulation of complement activity
Translation
Cytoplasmic translational initiation
0.000
0.011
0.102
0.030
0.097
0.149
0.007
0.011
0.005
0.007
0.083
0.123
0.506
0.645
Normalized enrichment score (q value shown)
MYCN non-amplified (13 patients)
All patients (14 patients)
