## Supplementary Figures S1-S6 and Supplementary Tables S1-S11 for "CLEC10A marks an MHC class II-high tumour myeloid state that lies in T-cell-rich neighbourhoods, and its survival association is seen in MYCN non-amplified neuroblastoma": Supplementary_Figure_S6.pdf

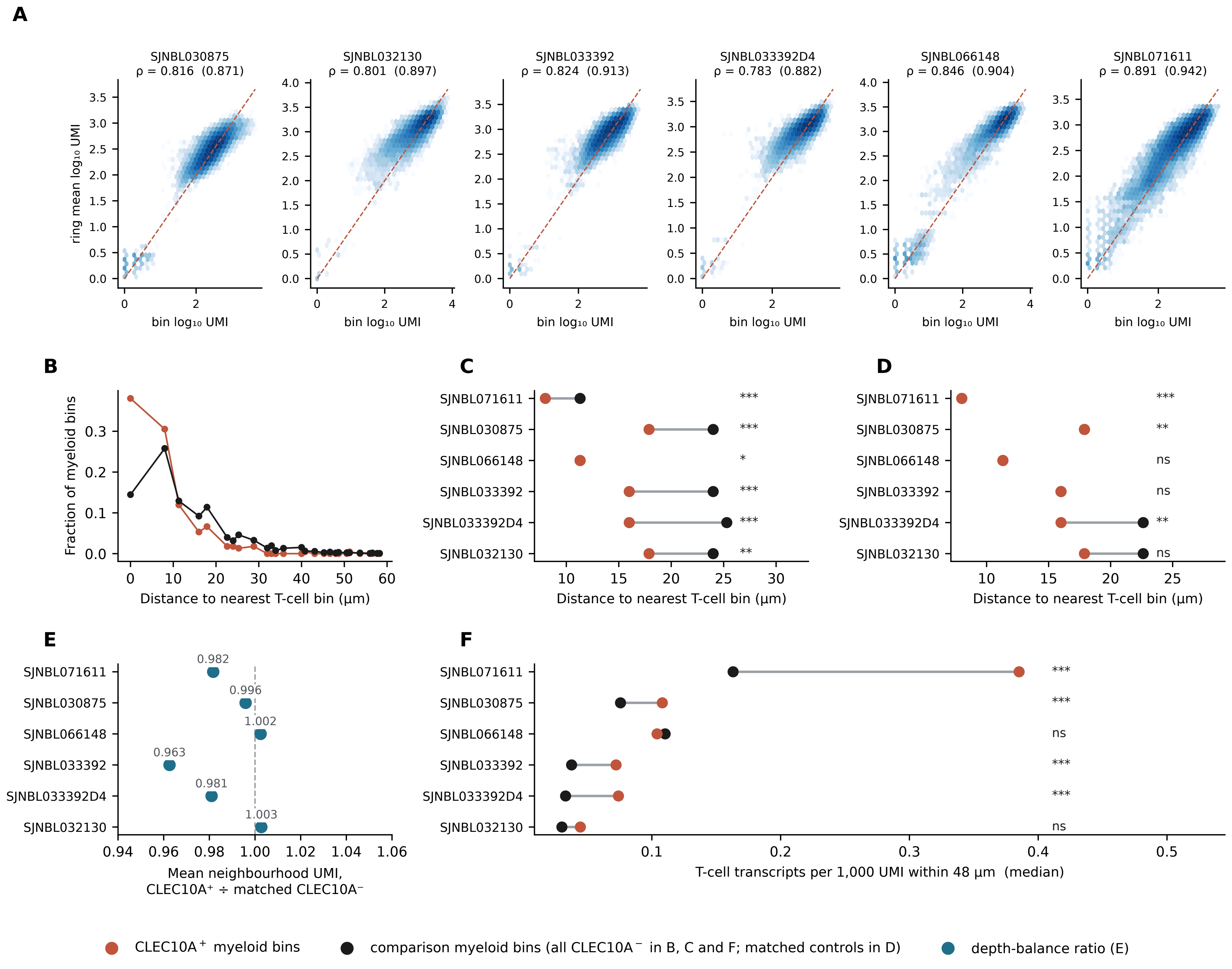

A
SJNBL030875
ρ = 0.816 (0.871)
SJNBL032130
ρ = 0.801 (0.897)
SJNBL033392
ρ = 0.824 (0.913)
SJNBL033392D4
ρ = 0.783 (0.882)
SJNBL066148
ρ = 0.846 (0.904)
SJNBL071611
ρ = 0.891 (0.942)
ring mean log₁₀ UMI
bin log₁₀ UMI
B
Fraction of myeloid bins
Distance to nearest T-cell bin (µm)
C
SJNBL071611 ***
SJNBL030875 ***
SJNBL066148 *
SJNBL033392 ***
SJNBL033392D4 ***
SJNBL032130 **
Distance to nearest T-cell bin (µm)
D
SJNBL071611 ***
SJNBL030875 **
SJNBL066148 ns
SJNBL033392 ns
SJNBL033392D4 **
SJNBL032130 ns
Distance to nearest T-cell bin (µm)
E
SJNBL071611 0.982
SJNBL030875 0.996
SJNBL066148 1.002
SJNBL033392 0.963
SJNBL033392D4 0.981
SJNBL032130 1.003
Mean neighbourhood UMI, CLEC10A⁺ ÷ matched CLEC10A⁻
F
SJNBL071611 ***
SJNBL030875 ***
SJNBL066148 ns
SJNBL033392 ***
SJNBL033392D4 ***
SJNBL032130 ns
T-cell transcripts per 1,000 UMI within 48 µm (median)
CLEC10A⁺ myeloid bins
comparison myeloid bins (all CLEC10A⁻ in B, C and F; matched controls in D)
depth-balance ratio (E)
